# Common variants at 21q22.3 locus influence *MX1* gene expression and susceptibility to severe COVID-19

**DOI:** 10.1101/2020.12.18.20248470

**Authors:** Immacolata Andolfo, Roberta Russo, Alessandro Vito Lasorsa, Sueva Cantalupo, Barbara Eleni Rosato, Ferdinando Bonfiglio, Giulia Frisso, Abete Pasquale, Gian Marco Cassese, Giuseppe Servillo, Gabriella Esposito, Ivan Gentile, Carmelo Piscopo, Romolo Villani, Giuseppe Fiorentino, Pellegrino Cerino, Carlo Buonerba, Biancamaria Pierri, Massimo Zollo, Achille Iolascon, Mario Capasso

## Abstract

The COVID-19 disease, caused by the SARS-Cov-2, presents a heterogeneous clinical spectrum. The risk factors do not fully explain the wide spectrum of disease manifestations, so it is possible that genetic factors could account for novel insights into its pathogenesis.

In our previous study, we hypothesized that common variants on chromosome 21, near *TMPRSS2* and *MX1* genes, may be genetic risk factors associated to the different clinical manifestations of COVID-19. Here, we performed an in-depth genetic analysis of chromosome 21 exploiting the genome-wide association study data including 6,406 individuals hospitalized for COVID-19 and 902,088 controls with European genetic ancestry from COVID-19 Host Genetics Initiative. We found that five single nucleotide polymorphisms (SNPs) within *TMPRSS2* and near *MX1* gene show suggestive associations (P≤1×10^−5^) with severe COVID-19. All five SNPs replicated the association in two independent cohorts of Asian subjects while two and one out of the 5 SNPs replicated in African and Italian populations, respectively (P≤0.05). The minor alleles of these five SNPs correlated with a reduced risk of developing severe COVID-19 and increased level of *MX1* expression in blood.

Our findings provide further evidence that host genetic factors can contribute to determine the different clinical presentations of COVID-19 and that MX1, an antiviral effector of type I and III interferon pathway, may be a potential therapeutic target.

## Introduction

The recent SARS-Cov-2 pandemic has caused so far more than one million of deaths (https://covid19.who.int/). The COVID-19, caused by the SARS-Cov-2, has several clinical conditions ranging from no or mild symptoms to rapid progression to respiratory failure (1, 2). Advanced age is a major risk factor, as well as male sex and some co-morbidities such as hypertension and diabetes (3). Since clinical risk factors do not fully explain the wide spectrum of disease manifestations, it is conceivable that dissecting the genetics of the disease may provide novel insights into its pathogenesis.

A recent genome-wide association study (GWAS) identified two susceptibility loci of severe COVID-19: the first locus on chromosome 3 harbors multiple genes (*SLC6A20, LZFTL1, CCR9, CXCR6, XCR1, FYCO1*) that could be functionally implicated in COVID-19 pathology; the second on chromosome 9 that defines the ABO blood groups (4). Another study showed that inactivating rare mutations in genes belonging to type I interferon pathway predispose to life-threatening COVID-19 pneumonia (5). Other very recent papers reported the results from the analysis of two large independent GWASs that validated the two previous risk loci and found novel variants at chromosome 19p13.3, 12q24.13 and 21q22.1 associated with severe COVID-19 (6, 7).

In our previous opinion article, based on allele frequencies across different populations and expression quantitative loci (eQTLs) data, we hypothesized that common variants on chromosome 21 near *TMPRSS2* and *MX1* genes may be genetic risk factors associated to the COVID-19 different clinical manifestations (8). In this study, to further support our hypothesis, we exploited GWAS data from the COVID-19 Host Genetics Initiative (9) and performed an in-depth mapping of chromosome 21 using summary statistics where common variants at this chromosome were associated with severe COVID-19 at genome-wide significance level (P ≤ 5×10^−8^). Using the cohort of 908,494 subjects with European origins, we found 5 single nucleotide polymorphisms (SNPs) at the *TMPRSS2/MX1* locus showing suggestive association with the disease. All 5 SNPs replicated the association in two independent cohorts of Asian subjects whereas two SNPs confirmed the association in African and one SNP in Italian cohort. Significant eQTLs signals were found for *MX1* in blood.

## Methods

### GWAS

The summary statistics of chromosome 21 were obtained from the GWAS dataset “B2_ALL_eur_leave_23andme” deposited in the COVID-19 Host Genetics Initiative website (9). It includes 6,406 laboratory confirmed SARS-CoV-2 infection and hospitalized for COVID-19 cases and 902,088 controls from general population with European genetic ancestry.

### Replication

The summary statistics of the SNPs used for the replication study were retrieved from the GenOMICC study (6). Three independent cohorts of cases and controls with different ethnicity were available throughout GenOMMIC GWAS study (REF 6): 182 individuals from African ancestry (AFR), 149 from East Asian ancestry (EAS), 237 from South Asian ancestry (SAS). Moreover, 157 hospitalized COVID-19 cases and 1388 controls with Italian origins were typed fort the rs12329760 variant by TaqMan® SNP Genotyping (Applied Biosystems by Thermo Fisher Scientific).

### Definition of independent genome-wide associated loci

Using the 74 significant SNPs with P≤1×10^−5^ of chromosome 21, we defined three independent associated loci by the following computational process. The SNPs were first sorted according to their association P value. Then, the lead SNP, considered as the most significant SNP in a given genomic locus, was removed from this list and assigned to an independent locus together with all other SNPs which have a r^2^ value less than or equal to 0.01 with this SNP. This procedure was recursively applied to the remaining SNPs in the list, so that each SNP could be assigned to a locus and no SNPs were left in the original list.

### Assessment of the functional role of the SNPs

Candidate regulatory SNPs were explored by HaploReg (https://pubs.broadinstitute.org/mammals/haploreg/haploreg.php) (10). Prediction of the functional impact of 14 SNPs at *TMPRSS2/MX1* locus was assessed by Genome Wide Annotation of VAriants (GWAVA) tool (htts://www.sanger.ac.uk/sanger/StatGen_Gwava) (11) and by Combined Annotation Dependent Depletion (CADD) tool (https://cadd.gs.washington.edu/) (12). The distribution of the eQTLs was explored by Genotype Tissue Expression (GTEx) database (https://www.gtexportal.org/home).

### Statistical analysis

Allele frequencies were compared using the Chi-square test. A two-sided P≤.05 was considered statistically significant.

## Results

### *TMPRSS2*/*MX1* locus is associated with severe COVID-19

To prove that common variants at *TMPRSS2*/*MX1* (21q22.3) locus may affect the susceptibility to severe COVID-19 onset, we analyzed the summary statistics of a large available GWAS dataset released from COVID-19 Host Genetics Initiative. The database includes 6,406 hospitalized cases and 902,088 controls with European ancestry. The region on chromosome 21 appears to be significantly associated with severe COVID-19 at the genome-wide level (https://www.covid19hg.org/results/) (6). To investigate whether more than one association signal may exist at chromosome 21, we selected 74 SNPs showing a P≤1×10^−5^ and among these we identified 3 independent loci (**Supplementary Table 1**). The most significant signal was represented by rs13050728 (P=2.76×10^−12^, OR=0.83, **Figure 1a**) that maps within the *INFRA2* gene. The other two signals showed a suggestive significance level (P≤1×10^−5^) and were tagged by rs111783124 (P=2.39×10^−6^, OR=1.17, **Figure 1b**) and rs3787946 (P=2.73×10^−6^, OR=0.87, **Figure 1c**), respectively. The rs3787946 maps in an intronic region of *TMPRSS2* and the first closest gene was *MX1* (**Figure 1c**); herein we named this locus as “*TMPRSS2*/*MX1*”. An in-depth inspection of *TMPRSS2*/*MX1* locus showed that 13 SNPs were in LD with the lead rs3787946 (r^2^>0.8, **Table 1**) and that the 5 most significant SNPs (P-values ranged from 2.7×10^−6^ to 5.8×10^−6^, **Table 1**) were in strong LD with each other (r^2^>=0.90, **Supplementary Figure 1**). The other 9 SNPs showed an LD with the lead SNP rs3787946 ranging from 0.8 to 0.9 and P-values ranging from 6×10^−4^ to 0.04 (**Table 1**). We then sought to replicate the associations of the 14 SNPs in three independent cohorts of cases and controls of GenOMMIC GWAS (6) with non-European ancestry. All the 11 available SNPs replicated in the EAS population; the top 5 SNPs replicated in SAS population whereas two out five SNPs in the AFR one (**Table 1**). By using TaqMan assay, we typed the rs12329760 variant in 157 hospitalized COVID-19 and 1388 controls with Italian origins (**Supplementary Table 2)** collected in our Institute and confirmed the minor allele as protective factor against the aggressive form of disease (**Table 1**, OR=0.72, P=0.05).

**Table 1.** Associations of SNPs at *TMPRSS2/MX1* risk locus in linkage disequilibrium with the lead rs3787946 SNP in different populations and prioritization scores.

| RS Number | EA | OA | MAF | r<sup>2</sup> | OR | P_EUR | OR | P_EAS | OR | P_SAS | OR | P_AFR | OR | P_ITA | *Region score | *TSS score | ^Function | ^Score |
| --- | --- | --- | --- | --- | --- | --- | --- | --- | --- | --- | --- | --- | --- | --- | --- | --- | --- | --- |
| rs3787946 | C | G | 0.23 | 1.00 | 0.87 | **2.73E-06** | 0.63 | **0.026** | 0.71 | **0.02** | 0.74 | **0.07** | NA | NA | 0.16 | 0.29 | INTRONIC | 2 |
| rs9983330 | G | A | 0.23 | 0.91 | 0.88 | **3.12E-06** | 0.54 | **0.004** | 0.73 | **0.04** | 0.79 | 0.16 | NA | NA | 0.31 | 0.64 | REGULATORY | 4 |
| rs12329760 | T | C | 0.24 | 0.90 | 0.88 | **3.13E-06** | 0.64 | **0.029** | 0.76 | **0.08** | 0.78 | 0.14 | 0.72 | **0.059** | **0.32** | **0.41** | **MISSENSE** | **7** |
| rs2298661 | A | C | 0.23 | 0.99 | 0.88 | **4.51E-06** | 0.63 | **0.030** | 0.67 | **0.01** | 0.60 | **0.01** | NA | NA | 0.18 | 0.35 | INTRONIC | 2 |
| rs9985159 | T | C | 0.23 | 0.98 | 0.88 | **5.80E-06** | 0.61 | **0.018** | 0.75 | **0.06** | 0.98 | 0.89 | NA | NA | 0.16 | 0.46 | INTRONIC | 2 |
| rs2298660 | T | C | 0.20 | 0.82 | 0.88 | 0.001 | NA | NA | NA | NA | NA | NA | NA | NA | 0.12 | 0.28 | INTRONIC | 2 |
| rs7364088 | A | G | 0.26 | 0.84 | 0.91 | 0.002 | NA | NA | NA | NA | NA | NA | NA | NA | 0.19 | 0.23 | INTRONIC | 2 |
| rs2298663 | T | C | 0.25 | 0.87 | 1.08 | **0.005** | 1.49 | **0.052** | 1.12 | 0.40 | 0.94 | 0.66 | NA | NA | 0.26 | 0.37 | REGULATORY | 4 |
| rs2094881 | C | T | 0.25 | 0.87 | 1.08 | **0.005** | 1.47 | **0.058** | 1.10 | 0.47 | 0.93 | 0.60 | NA | NA | 0.29 | 0.26 | REGULATORY | 4 |
| rs8131649 | T | C | 0.25 | 0.85 | 0.92 | **0.007** | 0.64 | **0.035** | 0.90 | 0.46 | 1.01 | 0.93 | NA | NA | 0.26 | 0.35 | REGULATORY | 4 |
| rs8134203 | T | C | 0.26 | 0.85 | 1.08 | **0.007** | 1.49 | **0.058** | 1.09 | 0.54 | 0.91 | 0.50 | NA | NA | 0.26 | 0.41 | REGULATORY | 4 |
| rs8134216 | T | C | 0.26 | 0.85 | 1.08 | **0.007** | 1.54 | **0.038** | 1.11 | 0.43 | 0.91 | 0.49 | NA | NA | 0.28 | 0.4 | REGULATORY | 4 |
| rs2104810 | A | G | 0.26 | 0.85 | 1.08 | **0.008** | 1.54 | **0.040** | 1.10 | 0.47 | 0.90 | 0.48 | NA | NA | 0.23 | 0.35 | REGULATORY | 4 |
| rs8131648 | C | T | 0.26 | 0.85 | 1.07 | 0.036 | NA | NA | NA | NA | NA | NA | NA | NA | 0.33 | 0.42 | REGULATORY | 4 |

\*Scores from GWAVA predictor tool
^Scores from CADD predictor tool
In bold the SNPs that replicated in at least one cohort
EA: Effect Allele; OA: Other Allele
EUR: Europe; EAS: Est Asia; SAS: South Asia; AFR: Africa; ITA: Italia
MAF: minor allele frequency
OR: Odds Ratio

**Figure 1.**
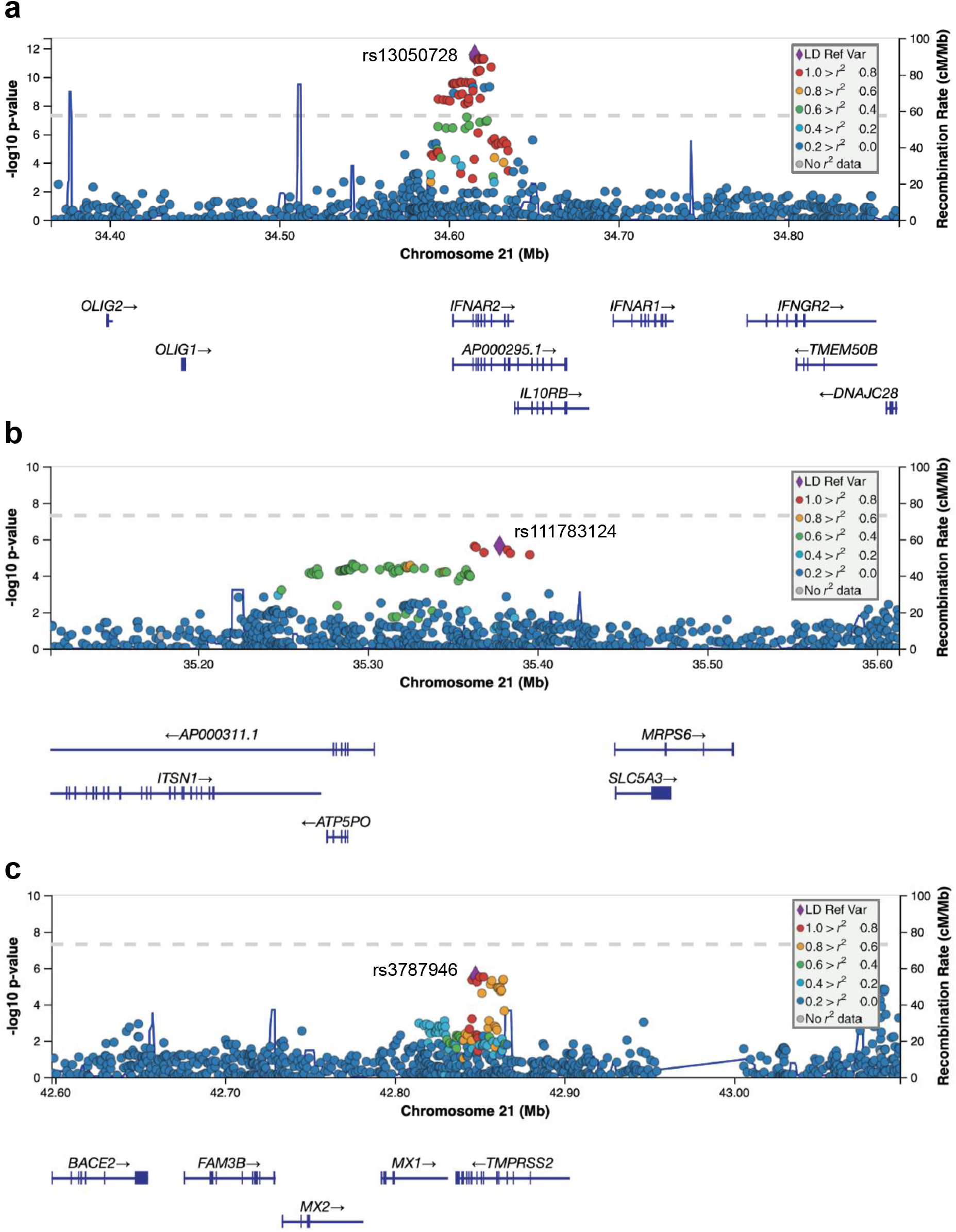
Regional association plots of the SNPs at three independent association signals of chromosome 21. Plots were generated using LocusZoom. Y□axes represent the significance of association (−log10 transformed P values) and the recombination rate. SNPs are color□coded based on pair□wise linkage disequilibrium (r^2^) with indicated lead SNPs: rs13050728 (panel a), rs111783124 (panel b) and rs3787946 (panel c).

### SNPs at *TMPRSS2*/*MX1* locus are enriched in regulatory regions active in thymus

We tested if the 14 SNPs (**Table 1**) and their proxy SNPs (r^2^>0.8) were significantly over- represented in active enhancers and promoters in multiple cell types and tissues by using HaploReg v4.1. These SNPs were enriched in the regulatory regions of several tissues (**Supplementary Table 3**), but the best enrichment was found in induced pluripotent stem cell and thymus (**Figure 2a**).

**Figure 2.**
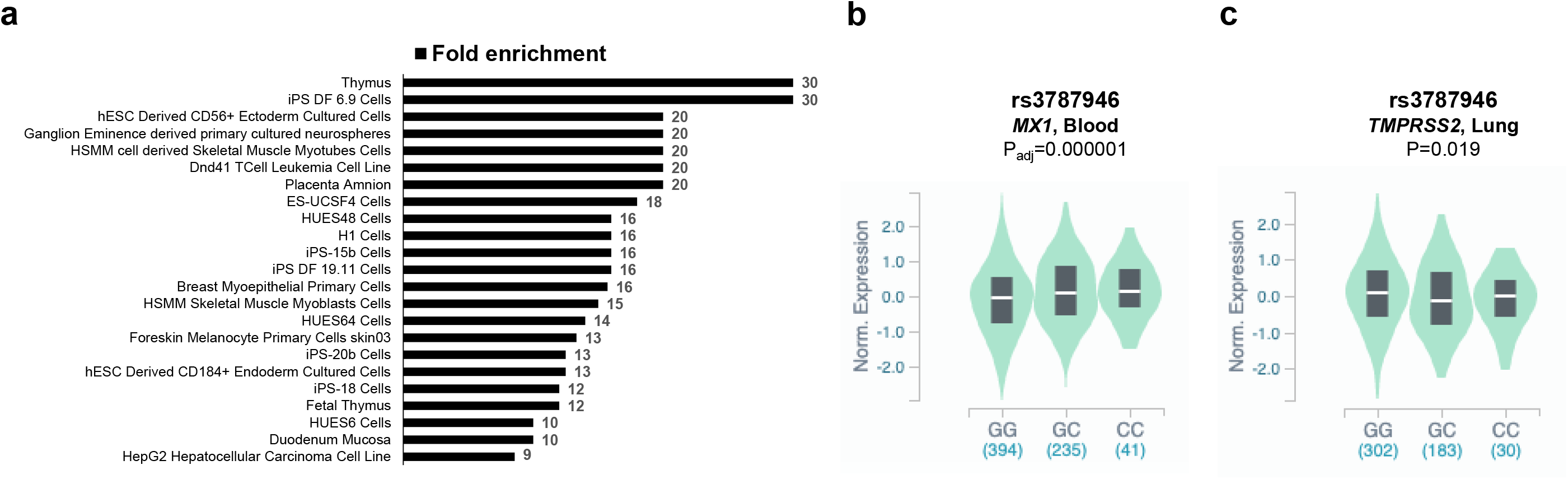
Enrichment of SNPs in regulatory regions and eQTL analyses. The statistically significant fold enrichments (P<0.05 after Bonferroni correction) of SNPs in regulatory DNA regions active in different tissues are shown (a). eQTL violin plots between genotypes of rs3787946 (b) and rs3787946 (c) with *MX1* and *TMPRSS2* expression from the from the Genotype-Tissue Expression (GTEx).

### Functional role of the most significant SNPs at *TMPRSS2*/*MX1* locus

We then investigated the predicted functional role of the 14 SNPs by GWAVA and CADD tools. We found that 2 (rs12329760 and rs9983330) out of 5 of the most significant SNPs showed the second and third most significant score (**Table 1**). The rs12329760 was classified as coding variant (p.Val197Met) localized in the exon 6 of *TMPRSS2* gene and was predicted to be pathogenic (PolyPhen=probably damaging and SIFT=deleterious).

### The most significant disease associated SNPs are eQTLs for *MX1* in blood

We verified if the top 5 SNPs (**Table 1**) might cause gene expression alterations interrogating the GTEx portal. We found that all the top 5 SNPs had eQTL signals for *MX1* exclusively in blood tissue. Particularly, the minor alleles of these SNPs correlated with higher expression of *MX1* compared to the major alleles (**Figure 2b, Supplementary Figure 2a**). Of note, all the 9 other SNPs, except for rs2298660, did not have eQTL signals for *MX1* in blood (**Supplementary Table 4**). The two SNPs, rs12329760 and rs2298660, were confirmed as eQTLs for *MX1* in blood (P=1.79×10^−6^ and 2.8×10^−6^, minor alleles correlated with a higher expression compared to the major alleles) by interrogation of another independent publicly available dataset (13). *TMPRSS2* is highly expressed in lung (8), so we investigated if the top 5 SNPs were eQTL for *TMPRSS2* in lung tissues at nominally statistically significant level (P≤0.05). We found that the minor alleles of 4 out 5 SNPs correlated with lower expression of *TMPRSS2* compared to the major alleles (**Figure 2c and Supplementary Figure 2b**). Notably, rs12329760 is also an eQTL for *TMPRSS2* in osteoblasts treated with dexamethasone (14).

## Discussion

Despite the substantial advances made in the recent months in the field of the SARS-CoV-2 infection, the major question remains about the identification of the factors that modulate the variable clinical spectrum of COVID-19.

Host genetic risk factors are emerging as a potential explanation for clinical heterogeneity of COVID-19 and are also crucial to find new druggable therapeutic targets (15). The main host cell entry factors of SARS-CoV-2 are ACE2 and TMPRSS2 (16, 17). The transmembrane spike (S) glycoprotein of virus binds to the ACE2 making it essential for the invasion of the virus into the host cell, followed by attachment of the virus to the target cells. S- protein priming by TMPRSS2 allows the binding of viral and cellular membranes, resulting in virus entry and replication in the host cells (18).

In our previous study, we hypothesized that common variants at chromosome 21, driving *TMPRSS2* and *MX1* expression, might have a mild-to-moderate effect in the susceptibility to SARS-CoV-2 infection. Particularly, genetic variants associated with reduced *TMPRSS2* and elevated *MX1* expression might confer less individual susceptibility to SARS-CoV-2 infection and favor a better outcome (8). Here, to further support our hypothesis, we exploited GWAS data of a cohort of 908,494 subjects with European origins from the COVID-19 Host Genetics Initiative (9) and performed an in-depth genetic analysis of chromosome 21. We identified five common variants (rs3787946, rs9983330, rs12329760, rs2298661 and rs9985159) at locus 21q22.3 within *TMPRSS2* and near *MX1* gene that showed suggestive associations (P≤1×10^−5^) with severe COVID-19. In particular, we found that the alleles with minor frequency were less recurrent among the hospitalized patients when compared to the control individuals, suggesting their protective role against the progression of disease. Interestingly, all the five SNPs replicated in two cohorts of Asian origin, whereas two SNPs replicated in a case series of African ancestry. Additionally, we replicated the association of the rs12329760 SNP in an independent case-control cohort of Italian origins. These results strongly indicate 21q22.3 as novel susceptibility locus to unfavorable outcome of COVID-19 and suggest that molecular mechanisms underlying this genetic predisposition may be common among individuals with different ethnicity.

Of note, the SNPs at this locus were enriched in the regulatory regions of induced pluripotent stem cell and thymus. Thymus plays a significant role in the regulation of adaptive immune responses. The effect of aging on the thymus and immune senescence is well established, and the resulting inflammaging is found to be implicated in the development of many chronic diseases. Both aging and diseases of inflammaging are associated with severe COVID-19, and a dysfunctional thymus may be implicated in the unfavorable outcome of disease (19, 20). Thus, our finding suggests that the identified SNPs could be severe COVID-19 risk factors also for their implications in the thymus function.

The five SNPs, here identified, had eQTL signals for *MX1* exclusively in blood tissue. Particularly, the minor allele of these SNPs correlated with higher expression of *MX1* and associated with a minor risk of developing severe COVID-19. These results support the evidence that *MX1* can play a relevant role in determining less severe forms of disease and are in line with a recent study that suggests *MX1* as antiviral effector against SARS-CoV-2 (21). Indeed, the expression of *MX1* was found to be higher in SARS-CoV-2 positive subjects, negatively correlated with age and independently associated with increased viral load (21). MX1 is part of the antiviral response induced by type I and III interferons (IFN) (22). Inactivating mutations in genes belonging to type I IFN pathway and the consequent decreased levels of proteins have been shown to recur in patients with severe COVID-19 (5). Therefore, our results further support the evidence that the use of drugs, activating IFN signaling, could be an effective treatment to prevent the adverse outcome of disease. There are already several ongoing clinical trials for COVID-19 prevention and/or treatment using type I or III IFNs (NCT04343976 [Phase II], NCT04385095 [Phase II], NCT04354259 [Phase II], NCT04293887 [Early Phase I], NCT04344600 [Phase II], NCT04320238 [Phase III], NCT04388709 [Phase II]). On the contrary, it is important to note that IFN administration could enhance a ‘‘cytokine-storm’’ causing a hyper-inflammatory response and contributing to multiple organ failure (21). Therefore, it is highly relevant to spawn new drugs that have the capacity to boost the host immune response while controlling tissue damage as a consequence of these infections.

We also report that the minor allele of 4 of the top 5 SNPs might reduce the expression of *TMPRSS2* in lung tissues and that the rs12329760 coding variant (p.Val197Met) correlate with lower expression of *TMPRSS2* in osteoblast treated with dexamethasone (14), a drug currently used to inhibit an excessive inflammation response (23). Of note, the rs12329760 coding variant was recently found to be less frequent among Chinese patients with critical COVID-19 disease (24). This variant is predicted to decrease the TMPRSS2 protein stability and ACE2 binding, thus decreasing the virus entry into the cells (25). Together, these data suggest that even the functions of *TMPRSS2* may be affected by risk variants of severe COVID-19. However, caution should be paid in considering these results as the eQTL signals in lung resulted to be not significant when corrected for multiple tests. Additional studies are needed to further verify the role of genetic variants *TMPRSS2*/*MX1* at locus in modulating the expression of *TMPRSS2*.

In conclusions, our results provide evidence that common variants, regulating the expression of *MX1*, can predispose to the risk of developing severe COVID-19. Unraveling the role of regulatory variants at the *TMPRSS2*/*MX1* locus could represent an important starting point for the treatment of COVID-19.

## Data Availability

Not applicable.

