## Supplemental material for "Common variants at 21q22.3 locus influence *MX1* gene expression and susceptibility to severe COVID-19"

**Article title:**

**Table of contents:**

- Table S1
- Table S2
- Table S3
- Table S4
- Figure S1
- Figure S2

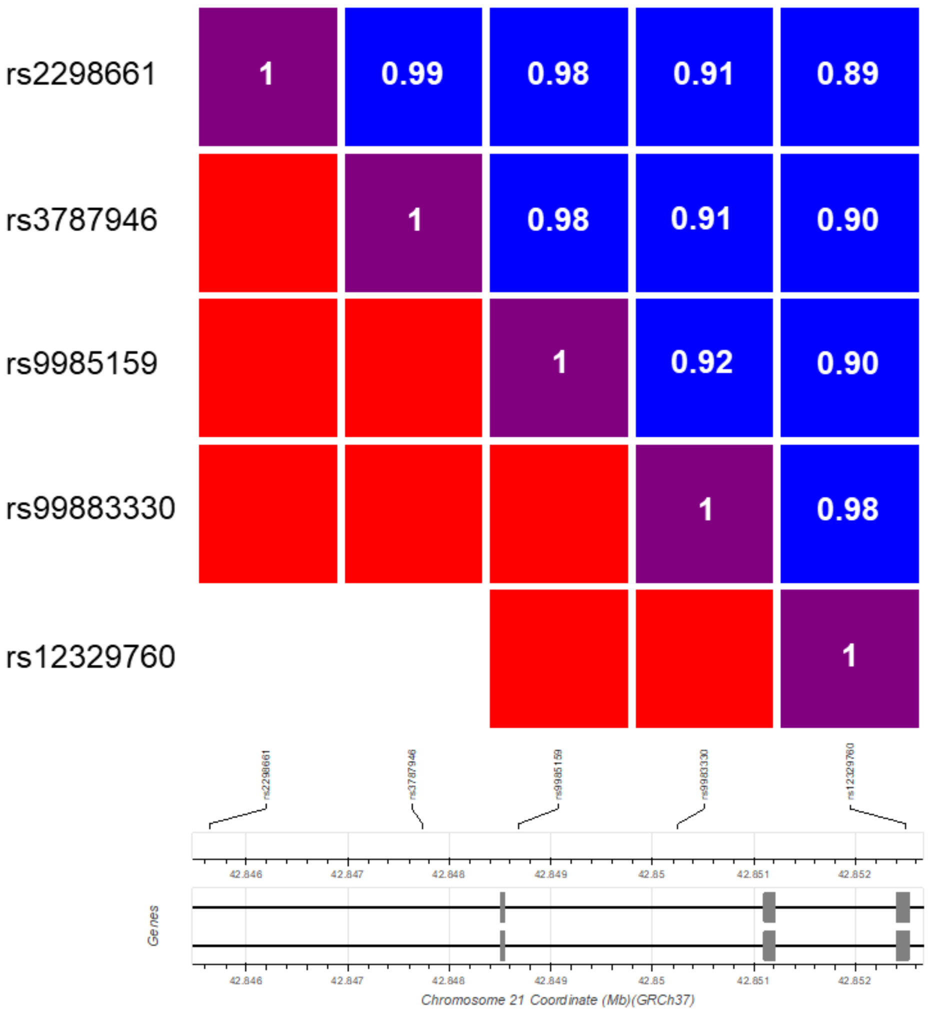

**Figure S1. Linkage disequilibrium block at *TMPRSS2/MX1* locus**

Linkage disequilibrium of the 5 most significant SNPs (P-values ranged from 2.7×10-6 to 5.8×10-6) with the lead rs3787946 at *TMPRSS2/MX1* locus. The D' and $r^2$ data are computed with the genetic information from European population by using the web tool LD-link (https://ldlink.nci.nih.gov/?tab=home).

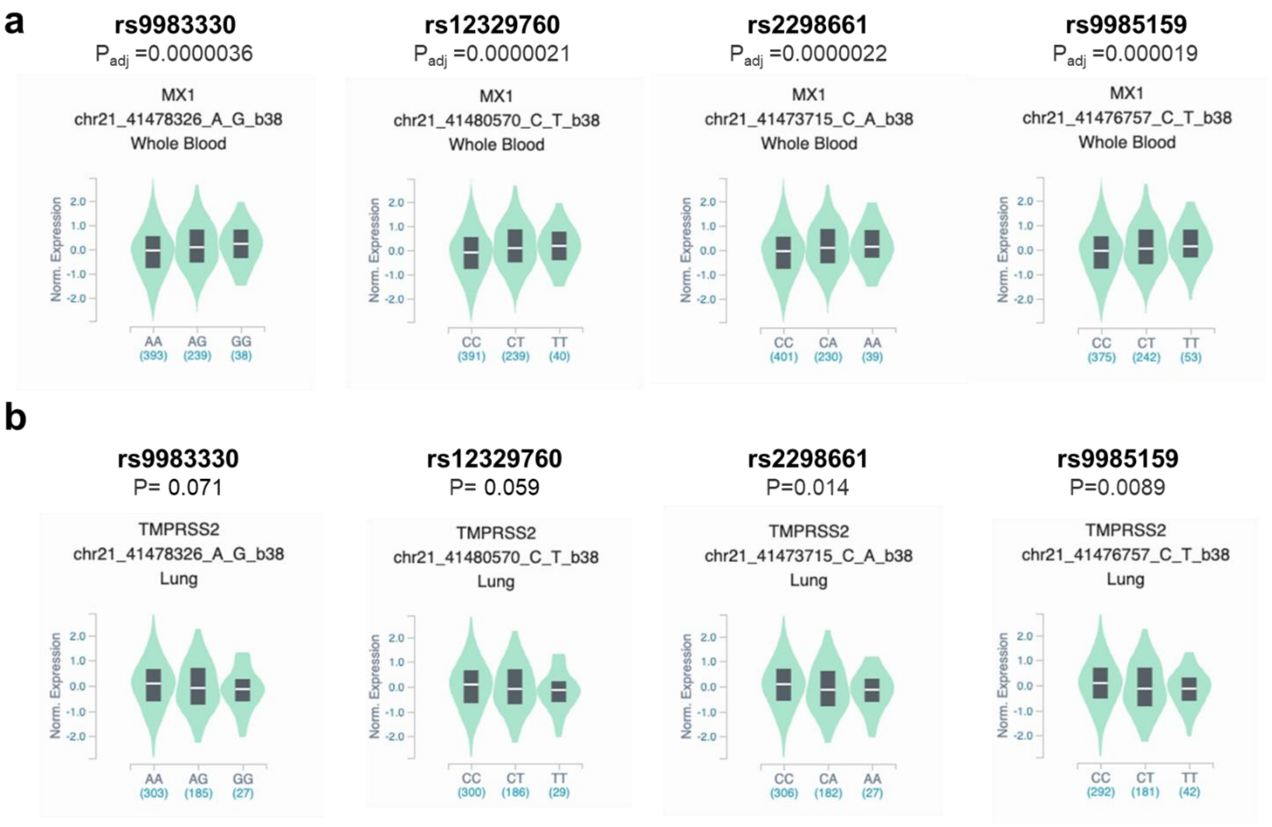

**Figure S1. Analysis of the eQTL signals of the top four disease associated SNPs in LD with the lead SNP rs3787946.**

Violin plots showing the eQTL signals for the rs9983330, rs12329760, rs2298661, and rs9985159 on *MX1* expression in whole blood (a) and on *TMPRSS2* expression in lung (b).

**Supplementary Table 1. Summary statistics at chromosome 21 from GWAS dataset (COVID-19 Host Genetics Initiative, "B2_ALL_eur_leave_23andme" )**

| CHR | POS | REF | ALT | SNP | all_meta_N | all_inv_var_meta_beta | all_inv_var_meta_sebeta | all_inv_var_meta_p | all_inv_var_het_p | all_meta_sample_N | all_meta_AF | rsid | OR | CI95L | CI95U |
|---|---|---|---|---|---|---|---|---|---|---|---|---|---|---|---|
| 21 | 3.5E+07 | T | C | 21:34615210:T:C | 13 | -1.82E-01 | 2.60E-02 | 2.76E-12 | 3.46E-02 | 905878 | 6.56E-01 | rs13050728 | 0.834 | 0.791 | 0.876 |
| 21 | 3.5E+07 | G | A | 21:34614834:G:A | 13 | -1.80E-01 | 2.60E-02 | 4.61E-12 | 3.53E-02 | 905878 | 6.56E-01 | rs9976829 | 0.835 | 0.793 | 0.878 |
| 21 | 3.5E+07 | A | G | 21:34617729:A:G | 13 | -1.80E-01 | 2.60E-02 | 4.98E-12 | 4.11E-02 | 905878 | 6.57E-01 | rs2252639 | 0.836 | 0.793 | 0.878 |
| 21 | 3.5E+07 | G | A | 21:34620451:G:A | 13 | -1.80E-01 | 2.60E-02 | 5.18E-12 | 3.23E-02 | 905878 | 6.57E-01 | rs2834163 | 0.836 | 0.793 | 0.878 |
| 21 | 3.5E+07 | A | G | 21:34619445:A:G | 13 | -1.80E-01 | 2.60E-02 | 5.38E-12 | 3.35E-02 | 905878 | 6.57E-01 | rs2073361 | 0.836 | 0.793 | 0.878 |
| 21 | 3.5E+07 | C | T | 21:34620207:C:T | 13 | -1.79E-01 | 2.60E-02 | 5.66E-12 | 3.15E-02 | 905878 | 6.57E-01 | rs2834161 | 0.836 | 0.793 | 0.879 |
| 21 | 3.5E+07 | C | A | 21:34616923:C:A | 13 | -1.79E-01 | 2.60E-02 | 5.90E-12 | 3.47E-02 | 905878 | 6.56E-01 | NA | 0.836 | 0.794 | 0.879 |
| 21 | 3.5E+07 | A | T | 21:34617950:A:T | 13 | -1.79E-01 | 2.60E-02 | 5.95E-12 | 5.02E-02 | 905878 | 6.57E-01 | rs2252650 | 0.836 | 0.793 | 0.879 |
| 21 | 3.5E+07 | A | G | 21:34624917:A:G | 13 | -1.79E-01 | 2.67E-02 | 2.01E-11 | 2.69E-02 | 905878 | 6.93E-01 | rs2236757 | 0.836 | 0.793 | 0.880 |
| 21 | 3.5E+07 | A | T | 21:34618043:A:T | 13 | -1.77E-01 | 2.66E-02 | 3.38E-11 | 3.41E-02 | 905878 | 6.93E-01 | rs2284549 | 0.838 | 0.794 | 0.882 |
| 21 | 3.5E+07 | A | G | 21:34618313:A:G | 13 | -1.76E-01 | 2.66E-02 | 3.53E-11 | 3.29E-02 | 905878 | 6.93E-01 | rs2284551 | 0.838 | 0.795 | 0.882 |
| 21 | 3.5E+07 | A | G | 21:34616545:A:G | 13 | -1.75E-01 | 2.66E-02 | 4.50E-11 | 2.85E-02 | 905878 | 6.93E-01 | rs2834157 | 0.839 | 0.796 | 0.883 |
| 21 | 3.5E+07 | A | C | 21:34606634:A:C | 13 | 1.65E-01 | 2.60E-02 | 2.22E-10 | 6.95E-02 | 905878 | 3.43E-01 | rs2834154 | 1.179 | 1.119 | 1.239 |
| 21 | 3.5E+07 | A | G | 21:34609944:A:G | 14 | 1.57E-01 | 2.48E-02 | 2.31E-10 | 7.43E-02 | 908494 | 3.43E-01 | rs9636867 | 1.170 | 1.113 | 1.227 |
| 21 | 3.5E+07 | G | A | 21:34607436:G:A | 13 | 1.65E-01 | 2.60E-02 | 2.35E-10 | 6.71E-02 | 905878 | 3.43E-01 | rs6517153 | 1.179 | 1.119 | 1.239 |
| 21 | 3.5E+07 | A | G | 21:34613301:A:G | 14 | 1.57E-01 | 2.47E-02 | 2.39E-10 | 8.60E-02 | 908494 | 3.43E-01 | rs17860169 | 1.170 | 1.113 | 1.226 |
| 21 | 3.5E+07 | C | G | 21:34611571:C:G | 13 | 1.58E-01 | 2.50E-02 | 2.58E-10 | 5.65E-02 | 634083 | 3.37E-01 | NA | 1.171 | 1.114 | 1.229 |
| 21 | 3.5E+07 | C | G | 21:34603249:C:G | 13 | 1.64E-01 | 2.60E-02 | 2.79E-10 | 5.72E-02 | 905878 | 3.43E-01 | NA | 1.178 | 1.118 | 1.238 |
| 21 | 3.5E+07 | G | A | 21:34604557:G:A | 13 | 1.64E-01 | 2.60E-02 | 2.87E-10 | 6.01E-02 | 905878 | 3.43E-01 | rs2300370 | 1.178 | 1.118 | 1.238 |
| 21 | 3.5E+07 | T | C | 21:34602934:T:C | 13 | 1.64E-01 | 2.60E-02 | 2.88E-10 | 5.30E-02 | 905878 | 3.43E-01 | rs12482556 | 1.178 | 1.118 | 1.238 |
| 21 | 3.5E+07 | C | A | 21:34602305:C:A | 13 | 1.64E-01 | 2.60E-02 | 3.01E-10 | 8.97E-02 | 905878 | 3.43E-01 | NA | 1.178 | 1.118 | 1.238 |
| 21 | 3.5E+07 | A | G | 21:34623919:A:G | 14 | 2.59E-01 | 4.17E-02 | 4.86E-10 | 9.05E-02 | 908494 | 7.73E-02 | rs17860220 | 1.296 | 1.190 | 1.402 |
| 21 | 3.5E+07 | T | C | 21:34614250:T:C | 14 | 2.53E-01 | 4.08E-02 | 5.19E-10 | 7.12E-02 | 908494 | 7.77E-02 | rs2229207 | 1.288 | 1.185 | 1.391 |
| 21 | 3.5E+07 | A | G | 21:34620801:A:G | 14 | 2.56E-01 | 4.14E-02 | 5.56E-10 | 8.71E-02 | 908494 | 7.81E-02 | rs2073362 | 1.292 | 1.188 | 1.397 |
| 21 | 3.5E+07 | T | G | 21:34614255:T:G | 13 | 1.52E-01 | 2.48E-02 | 9.15E-10 | 1.37E-01 | 907881 | 3.43E-01 | NA | 1.164 | 1.108 | 1.221 |
| 21 | 3.5E+07 | G | T | 21:34602794:G:T | 14 | 2.50E-01 | 4.12E-02 | 1.34E-09 | 4.25E-02 | 908494 | 7.74E-02 | rs17860118 | 1.284 | 1.180 | 1.388 |
| 21 | 3.5E+07 | C | G | 21:34607870:C:G | 13 | 1.61E-01 | 2.66E-02 | 1.53E-09 | 8.03E-02 | 905878 | 3.07E-01 | rs17860142 | 1.174 | 1.113 | 1.235 |
| 21 | 3.5E+07 | C | T | 21:34605778:C:T | 13 | 1.60E-01 | 2.66E-02 | 1.83E-09 | 8.61E-02 | 905878 | 3.07E-01 | rs2248420 | 1.174 | 1.112 | 1.235 |
| 21 | 3.5E+07 | G | A | 21:34618439:G:A | 14 | 1.50E-01 | 2.54E-02 | 3.17E-09 | 6.95E-02 | 908494 | 3.09E-01 | rs12053666 | 1.162 | 1.104 | 1.220 |
| 21 | 3.5E+07 | C | T | 21:34611730:C:T | 14 | 1.50E-01 | 2.54E-02 | 3.23E-09 | 7.79E-02 | 908494 | 3.06E-01 | rs17860165 | 1.162 | 1.104 | 1.220 |
| 21 | 3.5E+07 | T | A | 21:34593710:T:A | 13 | 1.57E-01 | 2.66E-02 | 3.53E-09 | 1.12E-01 | 905878 | 3.10E-01 | rs62226132 | 1.170 | 1.109 | 1.231 |
| 21 | 3.5E+07 | C | T | 21:34596750:C:T | 13 | 1.57E-01 | 2.66E-02 | 3.84E-09 | 9.87E-02 | 905878 | 3.10E-01 | rs62226152 | 1.170 | 1.109 | 1.231 |
| 21 | 3.5E+07 | G | A | 21:34599084:G:A | 13 | 1.56E-01 | 2.66E-02 | 4.17E-09 | 1.02E-01 | 905878 | 3.10E-01 | NA | 1.169 | 1.108 | 1.230 |
| 21 | 3.5E+07 | G | T | 21:34600508:G:T | 13 | 1.56E-01 | 2.66E-02 | 4.84E-09 | 9.04E-02 | 905878 | 3.09E-01 | NA | 1.168 | 1.108 | 1.229 |
| 21 | 3.5E+07 | C | T | 21:34611318:C:T | 14 | 1.48E-01 | 2.54E-02 | 5.76E-09 | 7.94E-02 | 908494 | 3.07E-01 | rs12482014 | 1.159 | 1.102 | 1.217 |
| 21 | 3.5E+07 | T | C | 21:34611545:T:C | 14 | 1.47E-01 | 2.53E-02 | 5.92E-09 | 5.32E-02 | 908494 | 3.07E-01 | rs12482193 | 1.159 | 1.101 | 1.216 |
| 21 | 3.5E+07 | G | A | 21:34609505:G:A | 13 | 1.48E-01 | 2.55E-02 | 7.50E-09 | 7.10E-02 | 907881 | 3.06E-01 | rs3153 | 1.159 | 1.101 | 1.217 |
| 21 | 3.5E+07 | T | C | 21:34610487:T:C | 12 | 1.34E-01 | 2.47E-02 | 6.37E-08 | 2.79E-01 | 905265 | 5.74E-01 | rs1131964 | 1.143 | 1.088 | 1.199 |
| 21 | 3.5E+07 | A | G | 21:34622536:A:G | 13 | -1.31E-01 | 2.48E-02 | 1.11E-07 | 2.66E-02 | 905878 | 4.86E-01 | rs2834165 | 0.877 | 0.834 | 0.919 |
| 21 | 3.5E+07 | A | C | 21:34621948:A:C | 13 | -1.31E-01 | 2.47E-02 | 1.26E-07 | 2.47E-02 | 905878 | 4.86E-01 | rs2834164 | 0.877 | 0.835 | 0.920 |
| 21 | 3.5E+07 | G | A | 21:34618285:G:A | 13 | -1.30E-01 | 2.47E-02 | 1.47E-07 | 1.98E-02 | 905878 | 4.88E-01 | rs2284550 | 0.878 | 0.836 | 0.921 |
| 21 | 3.5E+07 | A | G | 21:34611992:A:G | 12 | 1.29E-01 | 2.50E-02 | 2.42E-07 | 3.00E-01 | 667167 | 5.86E-01 | NA | 1.138 | 1.082 | 1.193 |
| 21 | 3.5E+07 | G | C | 21:34593574:G:C | 13 | 1.27E-01 | 2.47E-02 | 2.91E-07 | 1.76E-01 | 905878 | 5.67E-01 | NA | 1.135 | 1.080 | 1.190 |
| 21 | 3.5E+07 | T | G | 21:34602246:T:G | 11 | 1.28E-01 | 2.52E-02 | 3.72E-07 | 1.56E-01 | 392756 | 5.55E-01 | NA | 1.137 | 1.081 | 1.193 |
| 21 | 3.5E+07 | A | C | 21:34598385:A:C | 12 | 1.26E-01 | 2.48E-02 | 3.82E-07 | 1.38E-01 | 905265 | 5.67E-01 | rs1476415 | 1.134 | 1.079 | 1.189 |
| 21 | 3.5E+07 | T | G | 21:34609596:T:G | 12 | 1.26E-01 | 2.48E-02 | 4.00E-07 | 2.83E-01 | 905265 | 5.74E-01 | NA | 1.134 | 1.079 | 1.189 |
| 21 | 3.5E+07 | T | C | 21:34617213:T:C | 12 | -1.57E-01 | 3.18E-02 | 7.99E-07 | 3.58E-02 | 895822 | 6.56E-01 | rs2834158 | 0.855 | 0.801 | 0.908 |
| 21 | 3.5E+07 | C | T | 21:34626854:C:T | 12 | -1.52E-01 | 3.21E-02 | 2.04E-06 | 6.00E-02 | 895822 | 6.64E-01 | rs9975538 | 0.859 | 0.805 | 0.913 |

| | | | | | | | | | | | | | | | |
|---|---|---|---|---|---|---|---|---|---|---|---|---|---|---|---|
| 21 | 3.5E+07 | C | T | 21:35377591:C:T | 14 | 1.62E-01 | 3.43E-02 | 2.25E-06 | 1.81E-01 | 908494 | 1.29E-01 | NA | 1.176 | 1.097 | 1.255 |
| 21 | 3.5E+07 | G | A | 21:35362848:G:A | 14 | 1.56E-01 | 3.30E-02 | 2.39E-06 | 2.15E-01 | 908494 | 1.33E-01 | rs111783124 | 1.169 | 1.093 | 1.244 |
| 21 | 3.5E+07 | A | G | 21:34649337:A:G | 13 | 2.07E-01 | 4.40E-02 | 2.58E-06 | 2.73E-02 | 898438 | 1.25E-01 | NA | 1.230 | 1.124 | 1.336 |
| 21 | 4.3E+07 | G | C | 21:42847735:G:C | 14 | -1.34E-01 | 2.86E-02 | 2.73E-06 | 8.59E-01 | 908494 | 2.64E-01 | rs3787946 | 0.875 | 0.825 | 0.924 |
| 21 | 3.5E+07 | T | G | 21:35363759:T:G | 14 | 1.55E-01 | 3.30E-02 | 2.79E-06 | 2.15E-01 | 908494 | 1.33E-01 | rs11088268 | 1.167 | 1.092 | 1.243 |
| 21 | 3.5E+07 | A | G | 21:34625413:A:G | 12 | -1.49E-01 | 3.19E-02 | 2.90E-06 | 5.10E-02 | 895822 | 6.64E-01 | rs2236758 | 0.861 | 0.808 | 0.915 |
| 21 | 3.5E+07 | T | C | 21:34632316:T:C | 13 | 1.40E-01 | 2.99E-02 | 2.90E-06 | 1.71E-01 | 898438 | 3.36E-01 | rs2250226 | 1.150 | 1.083 | 1.217 |
| 21 | 4.3E+07 | A | G | 21:42850253:A:G | 14 | -1.32E-01 | 2.84E-02 | 3.12E-06 | 9.08E-01 | 908494 | 2.76E-01 | rs9983330 | 0.876 | 0.827 | 0.925 |
| 21 | 4.3E+07 | C | T | 21:42852497:C:T | 14 | -1.32E-01 | 2.83E-02 | 3.13E-06 | 8.87E-01 | 908494 | 2.75E-01 | rs12329760 | 0.876 | 0.828 | 0.925 |
| 21 | 3.9E+07 | A | C | 21:38577172:A:C | 8 | 7.37E-01 | 1.58E-01 | 3.29E-06 | 2.81E-01 | 380965 | 1.53E-02 | rs56309117 | 2.089 | 1.441 | 2.738 |
| 21 | 3.5E+07 | G | A | 21:35382261:G:A | 14 | 1.59E-01 | 3.44E-02 | 3.85E-06 | 2.81E-01 | 908494 | 1.26E-01 | NA | 1.172 | 1.093 | 1.251 |
| 21 | 4.3E+07 | C | T | 21:42864074:C:T | 14 | -1.23E-01 | 2.67E-02 | 4.25E-06 | 8.86E-01 | 908494 | 3.23E-01 | rs9305745 | 0.884 | 0.838 | 0.931 |
| 21 | 3.5E+07 | A | G | 21:34629175:A:G | 13 | 1.37E-01 | 2.98E-02 | 4.40E-06 | 2.11E-01 | 898438 | 3.36E-01 | rs11911133 | 1.147 | 1.080 | 1.214 |
| 21 | 3.5E+07 | T | C | 21:34592463:T:C | 14 | 2.64E-01 | 5.76E-02 | 4.45E-06 | 7.03E-01 | 908494 | 4.03E-02 | rs112268545 | 1.302 | 1.155 | 1.449 |
| 21 | 4.3E+07 | C | A | 21:42845642:C:A | 14 | -1.32E-01 | 2.87E-02 | 4.51E-06 | 8.81E-01 | 908494 | 2.64E-01 | NA | 0.877 | 0.827 | 0.926 |
| 21 | 3.5E+07 | A | G | 21:34631133:A:G | 13 | 1.36E-01 | 2.97E-02 | 4.62E-06 | 2.02E-01 | 898438 | 3.36E-01 | rs17860241 | 1.146 | 1.079 | 1.212 |
| 21 | 4.3E+07 | T | A | 21:42863723:T:A | 14 | -1.22E-01 | 2.67E-02 | 4.76E-06 | 9.23E-01 | 908494 | 3.22E-01 | rs10154090 | 0.885 | 0.839 | 0.931 |
| 21 | 3.5E+07 | C | G | 21:34634045:C:G | 12 | 1.47E-01 | 3.23E-02 | 4.94E-06 | 1.04E-01 | 895822 | 3.34E-01 | rs6517156 | 1.159 | 1.085 | 1.232 |
| 21 | 4.3E+07 | C | G | 21:42857322:C:G | 14 | -1.26E-01 | 2.75E-02 | 5.00E-06 | 9.18E-01 | 908494 | 3.10E-01 | rs9983252 | 0.882 | 0.834 | 0.929 |
| 21 | 3.5E+07 | G | A | 21:34590250:G:A | 14 | 2.60E-01 | 5.71E-02 | 5.25E-06 | 7.12E-01 | 908494 | 4.02E-02 | rs79997810 | 1.297 | 1.152 | 1.442 |
| 21 | 3.5E+07 | G | T | 21:35368402:G:T | 13 | 1.52E-01 | 3.35E-02 | 5.25E-06 | 1.42E-01 | 634083 | 1.25E-01 | rs12627254 | 1.165 | 1.088 | 1.241 |
| 21 | 3.5E+07 | C | G | 21:34627774:C:G | 11 | -1.48E-01 | 3.27E-02 | 5.57E-06 | 9.49E-02 | 621411 | 6.72E-01 | rs11701402 | 0.862 | 0.807 | 0.917 |
| 21 | 4.3E+07 | C | T | 21:42848684:C:T | 14 | -1.29E-01 | 2.85E-02 | 5.80E-06 | 9.14E-01 | 908494 | 2.64E-01 | NA | 0.879 | 0.829 | 0.928 |
| 21 | 3.5E+07 | A | G | 21:35383937:A:G | 14 | 1.55E-01 | 3.43E-02 | 5.83E-06 | 3.00E-01 | 908494 | 1.27E-01 | rs11088269 | 1.168 | 1.090 | 1.247 |
| 21 | 3.5E+07 | G | C | 21:35395439:G:C | 14 | 1.55E-01 | 3.45E-02 | 7.06E-06 | 2.50E-01 | 908494 | 1.24E-01 | rs11702497 | 1.168 | 1.089 | 1.247 |
| 21 | 4.3E+07 | T | C | 21:42856544:T:C | 14 | -1.23E-01 | 2.75E-02 | 7.80E-06 | 9.20E-01 | 908494 | 3.11E-01 | rs2838039 | 0.884 | 0.837 | 0.932 |

*The lead SNPs of the 3 indipendent locus are colored  in yellow*

**Supplementary Table 2. Patients' characteristics**

| Characteristic | Severe cases | % |
|---|---|---|
| | **N=157** | |
| **Age** | | |
| Years, mean (standard deviation) | 60.6 (17.2) | |
| Unknown | 2 | |
| **Sex - no. (%)** | | |
| Male | 92 | 59.4 |
| Female | 63 | 40.6 |
| **Previous coexisting disease - no. (%)** | | |
| 0-2 | 103 | 66.4 |
| >=3 | 26 | 16.8 |
| Unknown | 28 | 16.8 |

**Supplementary Table 3. Results of SNP enrichment analysis in regulotary elements in different tissues and cell types**

| Cell | Observed | Expected | Fold | Binomial p | ^adjusted_P |
|---|---|---|---|---|---|
| E112 THYM (Thymus) | 6 | 0.2 | 30.0 | 0 | 0 |
| E021 IPSC.DF.6.9 (iPS DF 6.9 Cells) | 6 | 0.2 | 30.0 | 0 | 0 |
| E012 ESDR.CD56.ECTO (hESC Derived CD56+ Ectoderm Cultured Cells) | 8 | 0.4 | 20.0 | 0 | 0 |
| E054 BRN.GANGEM.DR.NRSPHR (Ganglion Eminence derived primary cultured neurospheres) | 8 | 0.4 | 20.0 | 0 | 0 |
| E099 PLCNT.AMN (Placenta Amnion) | 6 | 0.3 | 20.0 | 0 | 0 |
| E115 BLD.DND41.CNCR (Dnd41 TCell Leukemia Cell Line) | 6 | 0.3 | 20.0 | 0 | 0 |
| E121 MUS.HSMMT (HSMM cell derived Skeletal Muscle Myotubes Cells) | 8 | 0.4 | 20.0 | 0 | 0 |
| E024 ESC.4STAR (ES-UCSF4 Cells) | 9 | 0.5 | 18.0 | 0 | 0 |
| E014 ESC.HUES48 (HUES48 Cells) | 8 | 0.5 | 16.0 | 0 | 0 |
| E003 ESC.H1 (H1 Cells) | 8 | 0.5 | 16.0 | 0 | 0 |
| E018 IPSC.15b (iPS-15b Cells) | 8 | 0.5 | 16.0 | 0 | 0 |
| E022 IPSC.DF.19.11 (iPS DF 19.11 Cells) | 8 | 0.5 | 16.0 | 0 | 0 |
| E027 BRST.MYO (Breast Myoepithelial Primary Cells) | 11 | 0.7 | 15.7 | 0 | 0 |
| E120 MUS.HSMM (HSMM Skeletal Muscle Myoblasts Cells) | 6 | 0.4 | 15.0 | 3.00E-06 | 0.000381 |
| E008 ESC.H9 (H9 Cells) | 3 | 0.2 | 15.0 | 0.001534 | 0.194818 |
| E016 ESC.HUES64 (HUES64 Cells) | 7 | 0.5 | 14.0 | 0 | 0 |
| E061 SKIN.PEN.FRSK.MEL.03 (Foreskin Melanocyte Primary Cells skin03) | 8 | 0.6 | 13.3 | 0 | 0 |
| E020 IPSC.20B (iPS-20b Cells) | 5 | 0.4 | 12.5 | 3.30E-05 | 0.004191 |
| E011 ESDR.CD184.ENDO (hESC Derived CD184+ Endoderm Cultured Cells) | 5 | 0.4 | 12.5 | 5.20E-05 | 0.006604 |
| E019 IPSC.18 (iPS-18 Cells) | 6 | 0.5 | 12.0 | 6.00E-06 | 0.000762 |
| E093 THYM.FET (Fetal Thymus) | 6 | 0.5 | 12.0 | 6.00E-06 | 0.000762 |
| E015 ESC.HUES6 (HUES6 Cells) | 6 | 0.6 | 10.0 | 9.00E-06 | 0.001143 |
| E077 GI.DUO.MUC (Duodenum Mucosa) | 4 | 0.4 | 10.0 | 0.000381 | 0.048387 |
| E098 PANC (Pancreas) | 4 | 0.4 | 10.0 | 0.000738 | 0.093726 |
| E094 GI.STMC.GAST (Gastric) | 3 | 0.3 | 10.0 | 0.002056 | 0.261112 |
| E007 ESDR.H1.NEUR.PROG (H1 Derived Neuronal Progenitor Cultured Cells) | 3 | 0.3 | 10.0 | 0.002349 | 0.298323 |
| E075 GI.CLN.MUC (Colonic Mucosa) | 2 | 0.2 | 10.0 | 0.01355 | 1.72085 |
| E101 GI.RECT.MUC.29 (Rectal Mucosa Donor 29) | 2 | 0.2 | 10.0 | 0.021315 | 2.707005 |
| E118 LIV.HEPG2.CNCR (HepG2 Hepatocellular Carcinoma Cell Line) | 6 | 0.7 | 8.6 | 3.20E-05 | 0.004064 |
| E074 BRN.SUB.NIG (Brain Substantia Nigra) | 3 | 0.4 | 7.5 | 0.008348 | 1.060196 |
| E059 SKIN.PEN.FRSK.MEL.01 (Foreskin Melanocyte Primary Cells skin01) | 2 | 0.3 | 6.7 | 0.044405 | 5.639435 |
| E090 MUS.LEG.FET (Fetal Muscle Leg) | 5 | 0.8 | 6.3 | 0.000715 | 0.090805 |
| E071 BRN.HIPP.MID (Brain Hippocampus Middle) | 3 | 0.5 | 6.0 | 0.011249 | 1.428623 |
| E053 BRN.CRTX.DR.NRSPHR (Cortex derived primary cultured neurospheres) | 3 | 0.5 | 6.0 | 0.014938 | 1.897126 |
| E001 ESC.I3 (ES-I3 Cells) | 3 | 0.5 | 6.0 | 0.014978 | 1.902206 |
| E088 LNG.FET (Fetal Lung) | 3 | 0.6 | 5.0 | 0.016702 | 2.121154 |
| E102 GI.RECT.MUC.31 (Rectal Mucosa Donor 31) | 2 | 0.4 | 5.0 | 0.049562 | 6.294374 |
| E013 ESDR.CD56.MESO (hESC Derived CD56+ Mesoderm Cultured Cells) | 2 | 0.4 | 5.0 | 0.071638 | 9.098026 |

| | | | | | |
|---|---|---|---|---|---|
| E002 ESC.WA7 (ES-WA7 Cells) | 1 | 0.2 | 5.0 | 0.175795 | 22.325965 |
| E109 GI.S.INT (Small Intestine) | 1 | 0.2 | 5.0 | 0.181152 | 23.006304 |
| E110 GI.STMC.MUC (Stomach Mucosa) | 2 | 0.5 | 4.0 | 0.085095 | 10.807065 |
| E066 LIV.ADLT (Liver) | 2 | 0.5 | 4.0 | 0.099955 | 12.694285 |
| E089 MUS.TRNK.FET (Fetal Muscle Trunk) | 2 | 0.6 | 3.3 | 0.119051 | 15.119477 |
| E070 BRN.GRM.MTRX (Brain Germinal Matrix) | 1 | 0.3 | 3.3 | 0.28264 | 35.89528 |
| E072 BRN.INF.TMP (Brain Inferior Temporal Lobe) | 1 | 0.4 | 2.5 | 0.322209 | 40.920543 |
| E068 BRN.ANT.CAUD (Brain Anterior Caudate) | 1 | 0.4 | 2.5 | 0.335088 | 42.556176 |
| E069 BRN.CING.GYR (Brain Cingulate Gyrus) | 1 | 0.4 | 2.5 | 0.337134 | 42.816018 |
| E116 BLD.GM12878 (GM12878 Lymphoblastoid Cells) | 1 | 0.4 | 2.5 | 0.343155 | 43.580685 |
| E026 STRM.MRW.MSC (Bone Marrow Derived Cultured Mesenchymal Stem Cells) | 1 | 0.5 | 2.0 | 0.394234 | 50.067718 |
| E005 ESDR.H1.BMP4.TROP (H1 BMP4 Derived Trophoblast Cultured Cells) | 1 | 0.5 | 2.0 | 0.40053 | 50.86731 |
| E084 GI.L.INT.FET (Fetal Intestine Large) | 1 | 0.5 | 2.0 | 0.404646 | 51.390042 |
| E129 BONE.OSTEO (Osteoblast Primary Cells) | 1 | 0.5 | 2.0 | 0.41587 | 52.81549 |
| E085 GI.S.INT.FET (Fetal Intestine Small) | 1 | 0.5 | 2.0 | 0.418745 | 53.180615 |
| E057 SKIN.PEN.FRSK.KER.02 (Foreskin Keratinocyte Primary Cells skin02) | 1 | 0.5 | 2.0 | 0.425214 | 54.002178 |
| E006 ESDR.H1.MSC (H1 Derived Mesenchymal Stem Cells) | 1 | 0.5 | 2.0 | 0.426514 | 54.167278 |
| E119 BRST.HMEC (HMEC Mammary Epithelial Primary Cells) | 1 | 0.6 | 1.7 | 0.449466 | 57.082182 |
| E028 BRST.HMEC.35 (Breast variant Human Mammary Epithelial Cells (vHMEC)) | 1 | 0.6 | 1.7 | 0.458849 | 58.273823 |
| E091 PLCNT.FET (Placenta) | 1 | 0.6 | 1.7 | 0.481212 | 61.113924 |
| E017 LNG.IMR90 (IMR90 fetal lung fibroblasts Cell Line) | 0 | 0.6 | 0.0 | 1 | 1 |
| E009 ESDR.H9.NEUR.PROG (H9 Derived Neuronal Progenitor Cultured Cells) | 0 | 0.4 | 0.0 | 1 | 1 |
| E010 ESDR.H9.NEUR (H9 Derived Neuron Cultured Cells) | 0 | 0.5 | 0.0 | 1 | 1 |
| E004 ESDR.H1.BMP4.MESO (H1 BMP4 Derived Mesendoderm Cultured Cells) | 0 | 0.3 | 0.0 | 1 | 1 |
| E062 BLD.PER.MONUC.PC (Primary mononuclear cells from peripheral blood) | 0 | 0.2 | 0.0 | 1 | 1 |
| E034 BLD.CD3.PPC (Primary T cells from peripheral blood) | 0 | 0.5 | 0.0 | 1 | 1 |
| E045 BLD.CD4.CD25I.CD127.TMEMPC (Primary T cells effector/memory enriched from peripheral bloo | 0 | 0.2 | 0.0 | 1 | 1 |
| E033 BLD.CD3.CPC (Primary T cells from cord blood) | 0 | 0.3 | 0.0 | 1 | 1 |
| E044 BLD.CD4.CD25.CD127M.TREGPC (Primary T regulatory cells from peripheral blood) | 0 | 0.3 | 0.0 | 1 | 1 |
| E043 BLD.CD4.CD25M.TPC (Primary T helper cells from peripheral blood) | 0 | 0.5 | 0.0 | 1 | 1 |
| E039 BLD.CD4.CD25M.CD45RA.NPC (Primary T helper naive cells from peripheral blood) | 0 | 0.4 | 0.0 | 1 | 1 |
| E041 BLD.CD4.CD25M.IL17M.PL.TPC (Primary T helper cells PMA-I stimulated) | 0 | 0.5 | 0.0 | 1 | 1 |
| E042 BLD.CD4.CD25M.IL17P.PL.TPC (Primary T helper 17 cells PMA-I stimulated) | 0 | 0.4 | 0.0 | 1 | 1 |
| E040 BLD.CD4.CD25M.CD45RO.MPC (Primary T helper memory cells from peripheral blood 1) | 0 | 0.4 | 0.0 | 1 | 1 |
| E037 BLD.CD4.MPC (Primary T helper memory cells from peripheral blood 2) | 0 | 0.5 | 0.0 | 1 | 1 |
| E048 BLD.CD8.MPC (Primary T CD8+ memory cells from peripheral blood) | 0 | 0.3 | 0.0 | 1 | 1 |
| E038 BLD.CD4.NPC (Primary T helper naive cells from peripheral blood) | 0 | 0.4 | 0.0 | 1 | 1 |
| E047 BLD.CD8.NPC (Primary T CD8+ naive cells from peripheral blood) | 0 | 0.4 | 0.0 | 1 | 1 |
| E029 BLD.CD14.PC (Primary monocytes from peripheral blood) | 0 | 0.6 | 0.0 | 1 | 1 |
| E031 BLD.CD19.CPC (Primary B cells from cord blood) | 0 | 0.4 | 0.0 | 1 | 1 |
| E035 BLD.CD34.PC (Primary hematopoietic stem cells) | 0 | 0.4 | 0.0 | 1 | 1 |
| E051 BLD.MOB.CD34.PC.M (Primary hematopoietic stem cells G-CSF-mobilized Male) | 0 | 0.6 | 0.0 | 1 | 1 |

| | | | | | |
|---|---|---|---|---|---|
| E050 BLD.MOB.CD34.PC.F (Primary hematopoietic stem cells G-CSF-mobilized Female) | 0 | 0.6 | 0.0 | 1 | 1 |
| E036 BLD.CD34.CC (Primary hematopoietic stem cells short term culture) | 0 | 0.5 | 0.0 | 1 | 1 |
| E032 BLD.CD19.PPC (Primary B cells from peripheral blood) | 0 | 0.5 | 0.0 | 1 | 1 |
| E046 BLD.CD56.PC (Primary Natural Killer cells from peripheral blood) | 0 | 0.5 | 0.0 | 1 | 1 |
| E030 BLD.CD15.PC (Primary neutrophils from peripheral blood) | 0 | 0.4 | 0.0 | 1 | 1 |
| E049 STRM.CHON.MRW.DR.MSC (Mesenchymal Stem Cell Derived Chondrocyte Cultured Cells) | 0 | 0.6 | 0.0 | 1 | 1 |
| E025 FAT.ADIP.DR.MSC (Adipose Derived Mesenchymal Stem Cell Cultured Cells) | 0 | 0.8 | 0.0 | 1 | 1 |
| E023 FAT.MSC.DR.ADIP (Mesenchymal Stem Cell Derived Adipocyte Cultured Cells) | 0 | 0.6 | 0.0 | 1 | 1 |
| E052 MUS.SAT (Muscle Satellite Cultured Cells) | 0 | 0.6 | 0.0 | 1 | 1 |
| E055 SKIN.PEN.FRSK.FIB.01 (Foreskin Fibroblast Primary Cells skin01) | 0 | 0.6 | 0.0 | 1 | 1 |
| E056 SKIN.PEN.FRSK.FIB.02 (Foreskin Fibroblast Primary Cells skin02) | 0 | 0.4 | 0.0 | 1 | 1 |
| E058 SKIN.PEN.FRSK.KER.03 (Foreskin Keratinocyte Primary Cells skin03) | 0 | 0.6 | 0.0 | 1 | 1 |
| E067 BRN.ANG.GYR (Brain Angular Gyrus) | 0 | 0.3 | 0.0 | 1 | 1 |
| E073 BRN.DL.PRFRNTL.CRTX (Brain Dorsolateral Prefrontal Cortex) | 0 | 0.3 | 0.0 | 1 | 1 |
| E082 BRN.FET.F (Fetal Brain Female) | 0 | 0.2 | 0.0 | 1 | 1 |
| E081 BRN.FET.M (Fetal Brain Male) | 0 | 0.5 | 0.0 | 1 | 1 |
| E063 FAT.ADIP.NUC (Adipose Nuclei) | 0 | 0.5 | 0.0 | 1 | 1 |
| E100 MUS.PSOAS (Psoas Muscle) | 0 | 0.3 | 0.0 | 1 | 1 |
| E108 MUS.SKLT.F (Skeletal Muscle Female) | 0 | 0.6 | 0.0 | 1 | 1 |
| E107 MUS.SKLT.M (Skeletal Muscle Male) | 0 | 0.6 | 0.0 | 1 | 1 |
| E083 HRT.FET (Fetal Heart) | 0 | 0.7 | 0.0 | 1 | 1 |
| E104 HRT.ATR.R (Right Atrium) | 0 | 0.4 | 0.0 | 1 | 1 |
| E095 HRT.VENT.L (Left Ventricle) | 0 | 0.5 | 0.0 | 1 | 1 |
| E105 HRT.VNT.R (Right Ventricle) | 0 | 0.4 | 0.0 | 1 | 1 |
| E065 VAS.AOR (Aorta) | 0 | 0.1 | 0.0 | 1 | 1 |
| E078 GI.DUO.SM.MUS (Duodenum Smooth Muscle) | 0 | 0.3 | 0.0 | 1 | 1 |
| E076 GI.CLN.SM.MUS (Colon Smooth Muscle) | 0 | 0.4 | 0.0 | 1 | 1 |
| E103 GI.RECT.SM.MUS (Rectal Smooth Muscle) | 0 | 0.3 | 0.0 | 1 | 1 |
| E111 GI.STMC.MUS (Stomach Smooth Muscle) | 0 | 0.3 | 0.0 | 1 | 1 |
| E092 GI.STMC.FET (Fetal Stomach) | 0 | 0.5 | 0.0 | 1 | 1 |
| E106 GI.CLN.SIG (Sigmoid Colon) | 0 | 0.3 | 0.0 | 1 | 1 |
| E079 GI.ESO (Esophagus) | 0 | 0.3 | 0.0 | 1 | 1 |
| E086 KID.FET (Fetal Kidney) | 0 | 0.2 | 0.0 | 1 | 1 |
| E097 OVRY (Ovary) | 0 | 0.4 | 0.0 | 1 | 1 |
| E087 PANC.ISLT (Pancreatic Islets) | 0 | 0.2 | 0.0 | 1 | 1 |
| E080 ADRL.GLND.FET (Fetal Adrenal Gland) | 0 | 0.7 | 0.0 | 1 | 1 |
| E096 LNG (Lung) | 0 | 0.3 | 0.0 | 1 | 1 |
| E113 SPLN (Spleen) | 0 | 0.4 | 0.0 | 1 | 1 |
| E114 LNG.A549.ETOH002.CNCR (A549 EtOH 0.02pct Lung Carcinoma Cell Line) | 0 | 0.4 | 0.0 | 1 | 1 |
| E117 CRVX.HELAS3.CNCR (HeLa-S3 Cervical Carcinoma Cell Line) | 0 | 0.4 | 0.0 | 1 | 1 |
| E122 VAS.HUVEC (HUVEC Umbilical Vein Endothelial Primary Cells) | 0 | 0.5 | 0.0 | 1 | 1 |
| E123 BLD.K562.CNCR (K562 Leukemia Cells) | 0 | 0.4 | 0.0 | 1 | 1 |

| | | | | | |
|---|---|---|---|---|---|
| E124 BLD.CD14.MONO (Monocytes-CD14+ RO01746 Primary Cells) | 0 | 0.4 | 0.0 | 1 | 1 |
| E125 BRN.NHA (NH-A Astrocytes Primary Cells) | 0 | 0.4 | 0.0 | 1 | 1 |
| E126 SKIN.NHDFAD (NHDF-Ad Adult Dermal Fibroblast Primary Cells) | 0 | 0.6 | 0.0 | 1 | 1 |
| E127 SKIN.NHEK (NHEK-Epidermal Keratinocyte Primary Cells) | 0 | 0.5 | 0.0 | 1 | 1 |
| E128 LNG.NHLF (NHLF Lung Fibroblast Primary Cells) | 0 | 0.4 | 0.0 | 1 | 1 |

*^P-values corrected according to Bonferroni method*

**Supplementary Table 4. Results of eQTL analysis**

| Gene | SNP | GWAS_P | eQTL_P | eQTL_P Threshold | *Statistically significant eQTL | NES | T-statistic | Tissue |
|---|---|---|---|---|---|---|---|---|
| MX1 | rs3787946 | 2.73E-06 | 0.0000011 | 0.000064 | YES | 0.17 | 4.9 | Whole Blood |
| MX1 | rs12329760 | 3.13E-06 | 0.0000021 | 0.000064 | YES | 0.17 | 4.8 | Whole Blood |
| MX1 | rs2298661 | 4.51E-06 | 0.0000022 | 0.000064 | YES | 0.17 | 4.8 | Whole Blood |
| MX1 | rs9983330 | 3.12E-06 | 0.0000036 | 0.000064 | YES | 0.16 | 4.7 | Whole Blood |
| MX1 | rs2298660 | 6.28E-04 | 0.0000140 | 0.000064 | YES | 0.15 | 4.4 | Whole Blood |
| MX1 | rs9985159 | 5.80E-06 | 0.0000190 | 0.000064 | YES | 0.15 | 4.3 | Whole Blood |
| MX1 | rs2094881 | 5.17E-03 | 0.0000660 | 0.000064 | 0 | -0.14 | -4 | Whole Blood |
| MX1 | rs7364088 | 2.27E-03 | 0.0000760 | 0.000064 | 0 | 0.13 | 4 | Whole Blood |
| MX1 | rs8131648 | 3.58E-02 | 0.0001100 | 0.000064 | 0 | -0.13 | -3.9 | Whole Blood |
| MX1 | rs8131649 | 6.55E-03 | 0.0001100 | 0.000064 | 0 | -0.13 | -3.9 | Whole Blood |
| MX1 | rs8134216 | 7.14E-03 | 0.0001300 | 0.000064 | 0 | -0.13 | -3.9 | Whole Blood |
| MX1 | rs8134203 | 7.10E-03 | 0.0001400 | 0.000064 | 0 | -0.13 | -3.8 | Whole Blood |
| MX1 | rs2298663 | 4.65E-03 | 0.0001600 | 0.000064 | 0 | -0.13 | -3.8 | Whole Blood |
| MX1 | rs2104810 | 7.86E-03 | 0.0007000 | 0.000064 | 0 | -0.12 | -3.4 | Whole Blood |

*Only SNPs with corrected P are considered statistically singifcant eQTLs
